# Long-read sequencing of the *ATP7B* gene from Moroccan patients with suspected Wilson disease

**DOI:** 10.1101/2025.10.13.25337866

**Authors:** Nada El Makhzen, Badreddine ElMakhzen, Laila Bouguenouch, Said Trhanint, Wafaa Bouzroud, Hakima Abid, Hugues Abriel

## Abstract

**Background:** Wilson disease (WD) is an autosomal recessive disease caused by loss of function of the copper transporter encoded by the *ATP7B gene*. Clinically, it mainly leads to hepatic failure and neuropsychiatric manifestations. Information about epidemiology, diagnosis, treatment, and survival of WD in Morocco is scarce. This study aimed to assess the feasibility of long-read sequencing using Oxford nanopore technology (ONT) of the coding and non-coding regions of *the ATP7B* gene in populations with limited access to genetic testing, such as the Moroccan population.

**Methods and Results:** We designed five long-range PCR primer pairs to amplify fragments approximately 15 kb long, encompassing the entire *ATP7B* gene. Barcoded libraries were prepared and sequenced via ONT flow cells (R10.4.1) using an Mk1C device. Our novel long-read ONT method was applied to 15 Moroccan patients clinically diagnosed with WD. This approach allowed the comprehensive detection of all single-nucleotide variants within the *ATP7B* gene. We identified 12 pathogenic variants, including a novel Alu insertion (c.1870-9_10ins) in intron 5, which is predicted to disrupt normal RNA processing and thereby cause WD. In all but one patient, the likely causative variants were identified, either in the homozygous or compound heterozygous state.

**Conclusion:** Our study demonstrates the versatility, affordability, and potential benefits of this methodology for the widespread implementation of *ATP7B* screening and diagnostic programs in WD patients from populations that have not yet been investigated. This is especially important for diseases that frequently exhibit heterozygote composites in genetically underexplored populations.

## 1. Introduction

Wilson disease (WD) (OMIM #277900) is an autosomal recessive genetic disease characterised by excessive copper accumulation in several organs, primarily the liver, brain, and eyes. It is caused by pathogenic variants in the *ATP7B* gene, which encodes a copper-transporting ATPase essential for controlling liver copper levels by leading copper to the secretory route and exporting excess copper into bile. If left untreated, WD can lead to life-threatening complications. However, with timely diagnosis and appropriate treatment, patients can significantly improve their health outcomes (1,2). WD prevalence ranges from 1/30,000 to 1/50,000 (3). The average age at WD diagnosis is 5–35 years (4). This disease is characterised as having an equal effect on men and women (5). Even though the clinical and genetic profiles of WD in North African individuals vary, there is a lack of comprehensive data on the disease in the Arab world, which is a significant concern due to the high prevalence of consanguinity (up to 69% depending on the region) (6). This can lead to a higher prevalence of homozygous variants of the *ATP7B* gene (7). Although specific data on the prevalence of WD in Morocco is limited, the high rate of consanguineous marriages in the country may contribute to a higher occurrence of this recessive disease. Additionally, WD poses both diagnostic and treatment challenges in Morocco. The diagnostic issue arises from the lack of genetic testing available in Moroccan facilities, which can often be too expensive in limited resource settings (8).

Integrating modern DNA sequencing technologies with appropriate bioinformatics pipelines could significantly reduce the time and costs involved in diagnosing rare diseases (9). Oxford Nanopore Technology (ONT) sequencing approach enables the detection of variants, including structural variants and tandem repeats, that are often overlooked by short-read sequencing methods. A recent study (10) demonstrated that ONT-based sequencing successfully identified diagnostic variants in previously undiagnosed cases of rare diseases, improving the accuracy and scope of genetic testing. This underscores ONT’s potential to increase diagnostic yield and provide a more comprehensive tool for clinical genomics in the diagnosis of rare diseases (10). However, LRS has its limitations and challenges compared with short-read NGS, including issues related to the quality of the initial samples and higher error rates. Additionally, the tools for raw data processing, mapping, and variant calling are less developed for LRS than for short-read NGS and are still being refined. Consequently, LRS has primarily been utilised thus far to investigate genetic diseases with known or highly suspected disease loci (9).

In this study, we assessed the feasibility and efficiency of using LRS from ONT to completely sequence the ∼80-kb *ATP7B* gene, encompassing both coding and non-coding regions, in individuals from the Moroccan population suspected of having WD.

## 2. Materials and methods

### 2.1. Recruitment of study participants and sample processing

Fifteen individuals were referred by hepatologists from the University Hospital Hassan II (Sidi Mohamed Ben Abdellah University, Fez, Morocco) to the Laboratory of Medical Genetics and Oncogenetics. Data were collected from the patients’ medical files, including their age at the onset of symptoms and at the time of diagnosis, their place of origin, and the presence of consanguinity. Additionally, we evaluated the results of various clinical examinations, including imaging and liver function tests (if performed), as well as the hepatic conditions that occurred throughout surveillance. Blood samples were then collected from those who consented to participate in the study (written informed consent was obtained from all patients or their legal guardians prior to genetic testing). This study protocol has been ethically approved by the appropriate institutional review board and is conducted in accordance with the ethical standards of the Declaration of Helsinki and relevant guidelines and regulations.

Samples were sent to the Medical Genetics Laboratory for DNA extraction and subsequently forwarded to the Ion Channels and Channelopathies Laboratory at the Institute for Biochemistry and Molecular Medicine (University of Bern) for long-read sequencing via Oxford Nanopore Technology (ONT).

### 2.2. Generation of sequence data

High-molecular-weight DNA was extracted from fresh whole-blood samples obtained from the University Hospital Hassan II via a Puregene DNA Extraction Kit (Gentra systems, Minneapolis, MN). For long-range PCR amplification, we used 100 ng/µL template DNA. We designed five overlapping long PCR products (Table 1), allowing the amplification of ∼15-kb fragments to cover the coding and noncoding regions of the *ATP7B* gene (Figure 1). The PCR mixture consisted of 12.5 µL of Takara Ex-Premier DNA Polymerase Dye plus (cat. #RR371A), 10.5 µL of nuclease-free H2O, and 1 μL of 10 pmol/μL forward and reverse primers (Table 1). Thermocycling comprised a 1-min denaturation step at 94°C, followed by 30 cycles at 98°C for 10 sec and 68°C for 13 min.

**Table 1:**
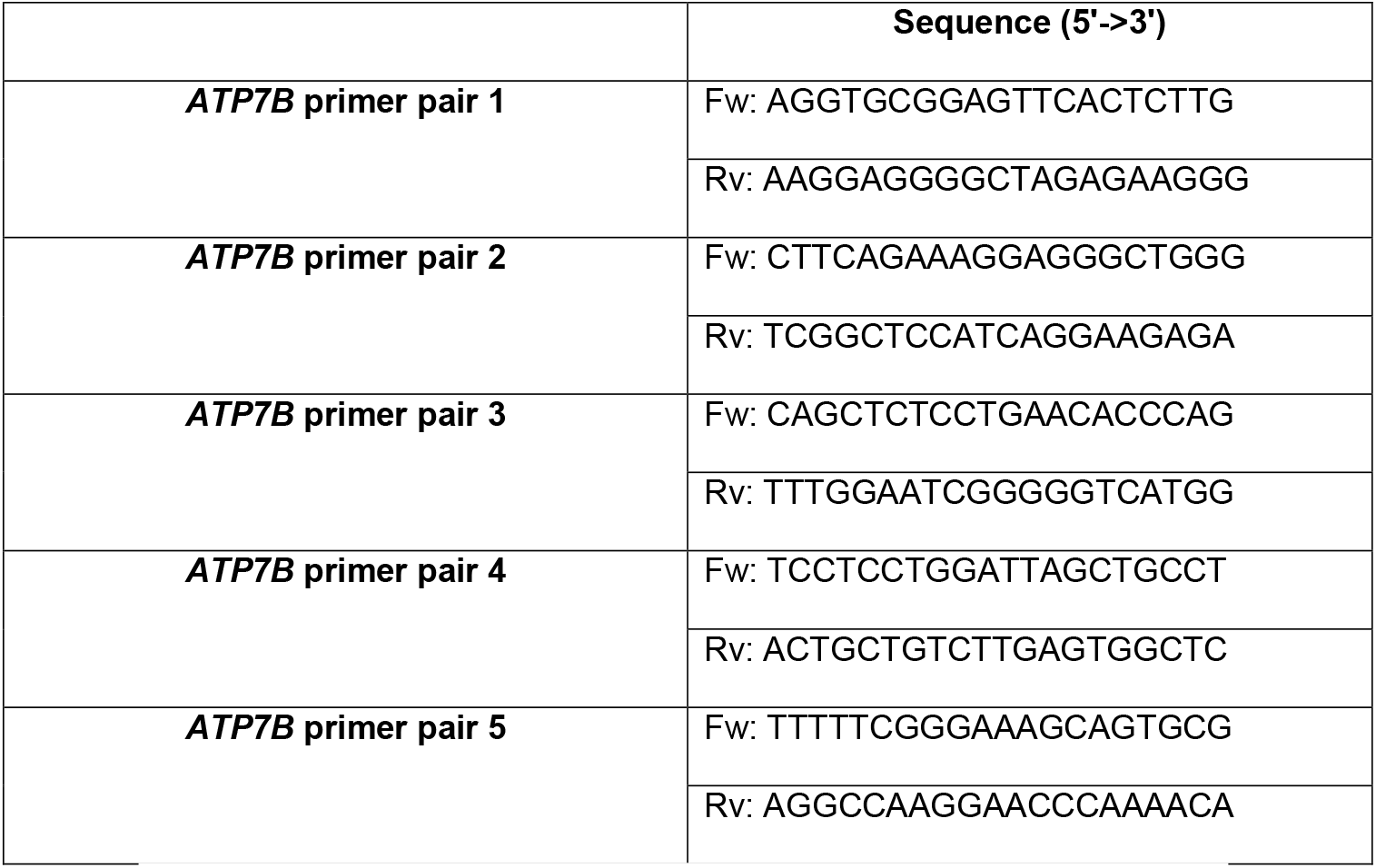
Primer pairs employed to generate the five amplicons of the *ATP7B* gene.

**Figure 1:**
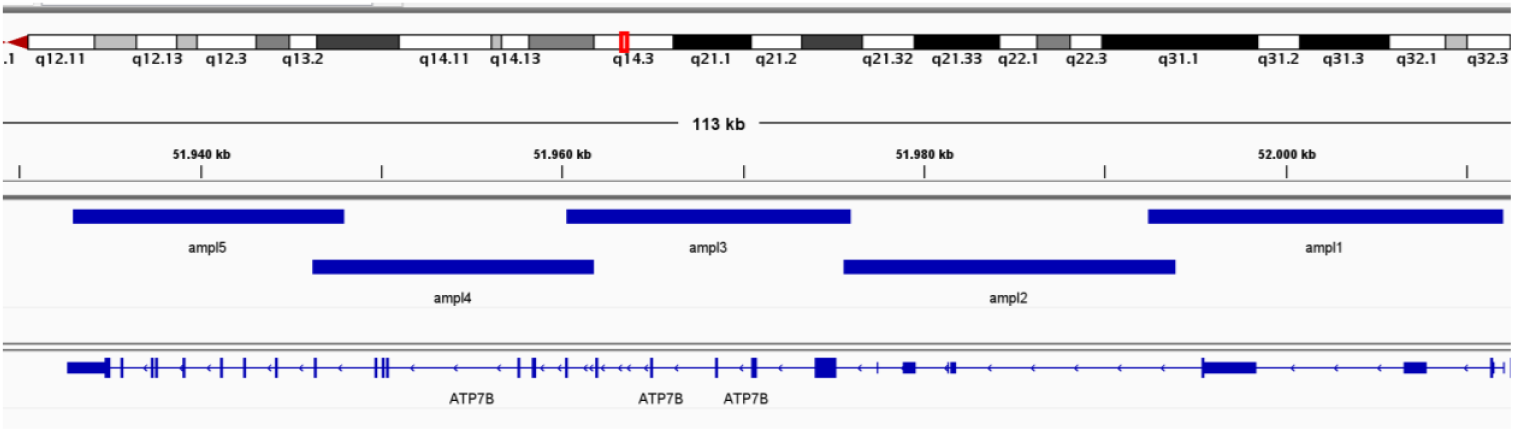
Illustration of the protocol for amplifying the ∼80-kb *ATP7B* gene via five long-range PCRs (amp.: amplicons), allowing the amplification of ∼15-kb fragments that encompass both the coding and noncoding regions of the *ATP7B* gene.

The amplified products were resolved on a 0.4% Tris-acetate EDTA agarose gel run at 160 V for 30 min. A D-DiGit Gel Scanner (LI-COR Biosciences, Homburg, Germany) was used to obtain images. The amplicons were subsequently purified with AMPure XP bead cleanup (Cat. #A63881) following the manufacturer’s protocol; the steps involved measuring the DNA volume and adding 1.8 volumes of AMPure XP beads to the sample. DNA fragments bound to paramagnetic beads, and DNA fragments plus beads were then separated from contaminants. The beads and DNA fragments were washed twice with 200 µL of 80% ethanol to remove impurities and eluted at the end with 40 µL of elution buffer (10 mM Tris-pH 8; 50 mM NaCl). The concentration of the purified PCR products was quantified via a NanoDrop OneC spectrophotometer (Thermo Scientific, MA, USA). For each patient, 50 ng of each purified PCR product (11 PCRs) was pooled and barcoded via the ONT Rapid Barcoding Kit 24 V14 (SQK-RBK114.24) following the manufacturer’s instructions. The pooled barcoded library of all samples was loaded onto one R10.4.1 flow cell (FLO-MIN114) and sequenced for 24 h on a MinION Mk1C sequencer.

### 2.3. Bioinformatics analysis

We converted the FAST5 output from sequencing into POD5 format and performed offline base-calling via Dorado v0.8.3 with the ‘sup’ model. We conducted quality control on the resulting FASTQ files via FastQC v0.12.1 and NanoPlot v1.41.6 (11). The ONT reads were filtered with fastp v0.23.4 to remove low-quality reads, defined as those with less than 40% of bases having a quality score below (12).

We aligned the filtered reads to the GRCh38 human reference genome via minimap2 v2.28 with the ‘lr:hq’ preset, which is optimized for Q20+ ONT reads (13). Mapping and depth statistics were calculated via the SAMtools v1.20 flagstat command and Mosdepth v0.3.8 (14,15). For variant calling, single-nucleotide variants (SNVs) and insertion/deletion variants (indels) were identified for each sample via Clair3 v1.0.10 (16) on the basis of the BAM files generated during the mapping process. The genomic VCF (gVCF) files produced in the variant calling step were then utilized for joint genotype calling across the entire cohort with GLnexus v1.4.1 (17), using the configuration file provided by Clair3.

We identified large structural variants (SVs), including an Alu insertion from the mapped ONT reads via Sniffles2 v2.4 (18) and cuteSV v2.1.1 (19). For Sniffles2, we first called SV candidates for each individual separately and then performed a population-level calling step across the cohort, using the SNF files from the initial step as input. For cuteSV, we called each individual separately following the recommended settings for ONT data and subsequently merged the sample-wise VCF files via SURVIVOR v1.0.7, applying a maximum breakpoint distance of 1 kb and a minimum support of 1, while accounting for the SV type and strand. We then proceeded to force-call the SVs in the merged VCF file for each individual via cuteSV in genotype mode, followed by another round of merging, resulting in sample-wise VCF files into a population-level VCF file with SURVIVOR.

To annotate the VCF files, we used the Ensembl Variant Effect Predictor (VEP) (20). The aligned reads were visualized via the Integrative Genomics Viewer (IGV). Variants with an allele frequency greater than 1% in public databases (gnomAD V4.1.0, dbSNP) were excluded from further analysis. The remaining variants were prioritized on the basis of their presence in various databases (ClinVar, HGMD Professional 2023.4), allele transmission (e.g., biallelic variants), and predicted severity. Candidate variants were assessed for deleterious effects via multiple in silico prediction tools, such as PolyPhen-2 and SIFT. It is important to note that phasing has not been performed for the variants included in this study. Variant classification was conducted according to the criteria established by the American College of Medical Genetics and Genomics (ACMG) (21).

### 2.4. Sanger sequencing

All *ATP7B* variants identified by ONT were subjected to Sanger sequencing for confirmation and segregation. Genomic DNA was used as a template to amplify the corresponding *ATP7B* exons/introns. Sequencing was performed via a 3500 Dx Genetic Analyser (Applied Biosystems, Foster City, CA, USA) with Big-Dye Terminator v3.1 chemistry (Applied Biosystems, Foster City, CA, USA). The resulting sequence data were analysed via a sequence software analyser.

### 2.5. MLPA

Detection of large deletions and duplications was performed using MLPA (Multiplex Ligation-dependent Probe Amplification) assay (SALSA MLPA P098 WD probemix, MRC-Holland, Amsterdam, Netherlands) in cases where only one or no variant were identified in the coding regions, adjacent splice site regions, and intronic regions.

## 3. Results

All 15 patients included in this study met the clinical diagnostic criteria for WD, scoring above 4 following the Leipzig 2001 criteria (EASL, 2012). The median age at diagnosis was 15 years. Among the patients, nine were from first-degree consanguineous marriages. Four patients presented with hepatic symptoms, including jaundice and ascites; two patients presented with neurological involvement; and four patients presented with visible Kayser–Fleischer rings on slit−lamp examination. Three patients were asymptomatic and were identified during a family screening survey, which revealed no functional or physical signs of the disease. Seven patients had liver involvement without any neurological symptoms. Notably, none of the patients displayed signs of cardiac, renal, or osteoarticular involvement. Biochemical evaluation revealed a disruption in copper homeostasis in the 15 patients (supplementary materials). Specifically, all patients presented decreased serum ceruloplasmin and copper levels, along with elevated 24-hour urinary copper excretion. These findings were consistent with a diagnosis of WD and supported the results of the clinical assessments.

We analysed the coding and noncoding regions of the complete *ATP7B* gene from 15 Moroccan individuals via ONT amplicon sequencing. In total, using the filtering approach described in “methods”, we identified 12 variants, including three that were classified as variants of uncertain significance (VUS), along with an insertion of an intronic Alu sequence (Table 2). Five patients presented with compound heterozygous variants, which aligns with the high frequency of compound variants. One patient was negative for both SNV variants and structural variants; MLPA results were normal (Figure 2).

**Table 2:**
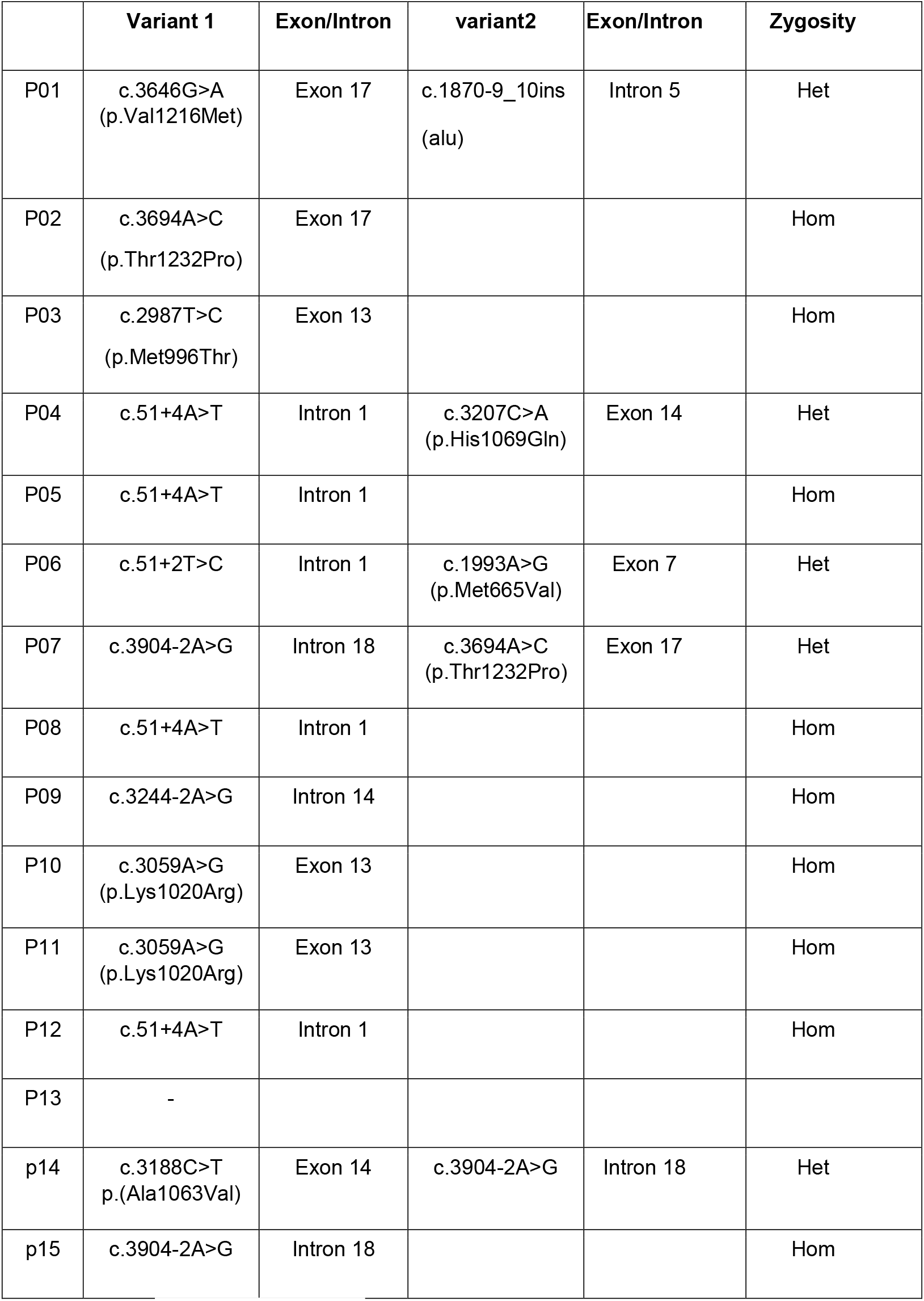
Identified *ATP7B* gene variants.

|  | Variant 1 | Exon/Intron | variant2 | Exon/Intron | Zygosity |
| --- | --- | --- | --- | --- | --- |
| P01 | c.3646G>A<br>(p.Val1216Met) | Exon 17 | c.1870-9_10ins<br>(alu) | Intron 5 | Het |
| P02 | c.3694A>C<br>(p.Thr1232Pro) | Exon 17 |  |  | Hom |
| P03 | c.2987T>C<br>(p.Met996Thr) | Exon 13 |  |  | Hom |
| P04 | c.51+4A>T | Intron 1 | c.3207C>A<br>(p.His1069Gln) | Exon 14 | Het |
| P05 | c.51+4A>T | Intron 1 |  |  | Hom |
| P06 | c.51+2T>C | Intron 1 | c.1993A>G<br>(p.Met665Val) | Exon 7 | Het |
| P07 | c.3904-2A>G | Intron 18 | c.3694A>C<br>(p.Thr1232Pro) | Exon 17 | Het |
| P08 | c.51+4A>T | Intron 1 |  |  | Hom |
| P09 | c.3244-2A>G | Intron 14 |  |  | Hom |
| P10 | c.3059A>G<br>(p.Lys1020Arg) | Exon 13 |  |  | Hom |
| P11 | c.3059A>G<br>(p.Lys1020Arg) | Exon 13 |  |  | Hom |
| P12 | c.51+4A>T | Intron 1 |  |  | Hom |
| P13 | - |  |  |  |  |
| p14 | c.3188C>T<br>p.(Ala1063Val) | Exon 14 | c.3904-2A>G | Intron 18 | Het |
| p15 | c.3904-2A>G | Intron 18 |  |  | Hom |

**Figure 2:**
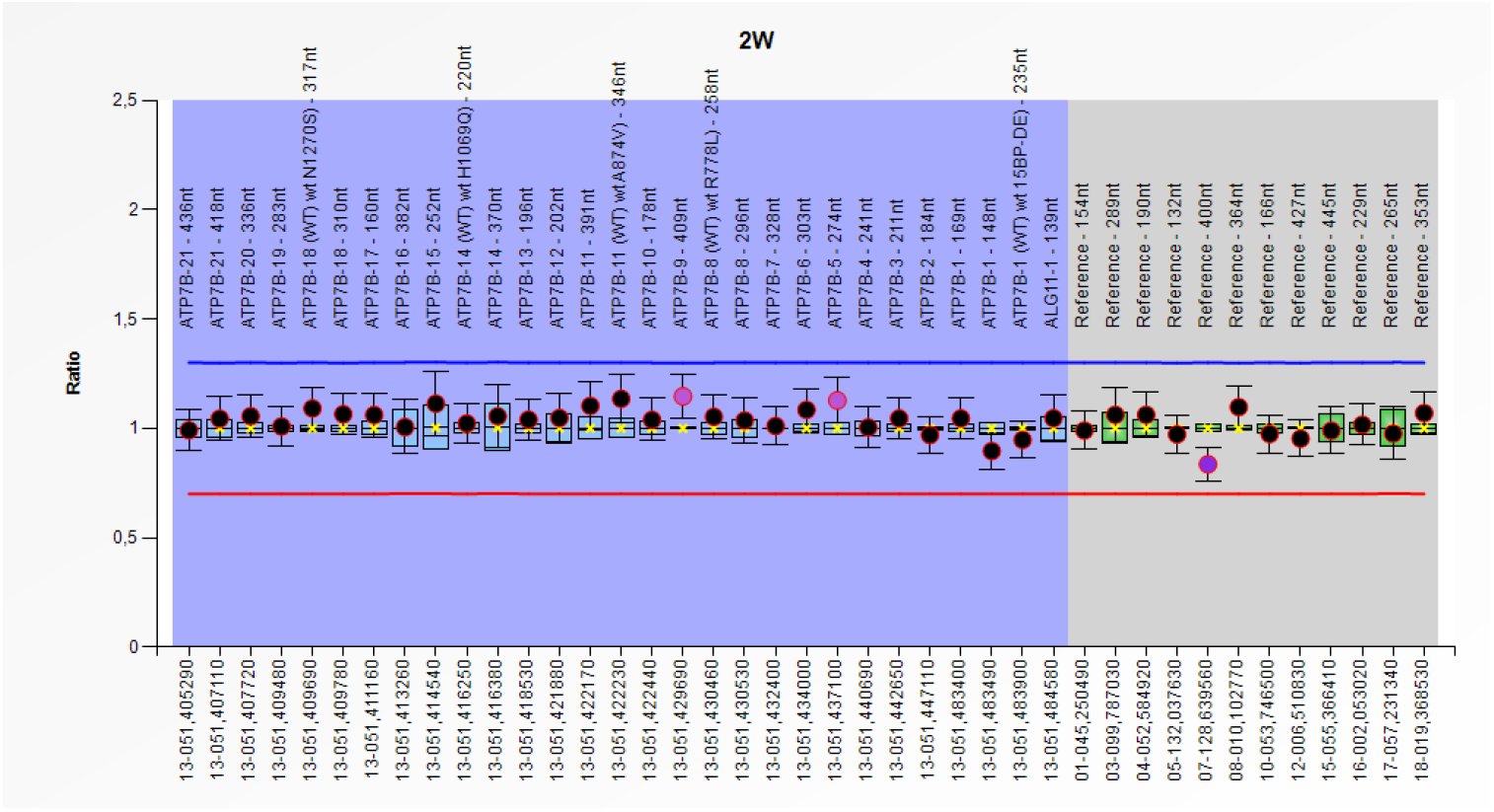
MLPA analysis of *ATP7B* in patient with normal results.

One patient in our cohort was heterozygous for a missense variant in exon 17, c.3646G>A (p.Val1216Met), which was inherited from her mother. This patient also had an insertion of an Alu sequence at position NC_000013.11:g.51961922_51961923ins (ATP7B): NM_000053.4:c.1870-10_1870-9ins[312]) in intron 5. (NM_000053.4:c.1870-10_1870-9insTTTTTTTTTTTTT GAGACGGAGTCTCGCTCTGTCGCCCAGGCCGGACTGGAGTACAGTGGCGCGATCTCGGCTCACTGCAAGCTCCGCTTCCCGGGTTCACGCCATTCTCCTGCCTCAGCCTCCCGAGTAGCTGGGACTACAGGCGCCCGCCACCACACGCCCGGCTAATTTTTTGTATTTTTAGTAGAGACGGGGTTTCACCTGTTAGCCAGGATGGTCTCGATCTCCTGACCTCATGATCCACCCGCCTCGGCCTCCCAAAGTGCTGGGATTACAGGCGTGAGCCACCGCGCCAGCCCACCCCCTCTCTTTT).

We designed primers to amplify the intronic region to confirm the presence of the insertion in a heterozygous state. The results from the agarose gel electrophoresis are shown in Figure 2, which illustrates two bands for the patient; one corresponds to the normal size of the amplicon (278 bp), whereas the second band is 650 bp in size (Figure 3). Sanger sequencing of the amplicon further confirmed the presence of a heterozygous insertion. Segregation analysis revealed that the insertion was absent in the mother. The father could not be tested as he is deceased.

**Figure 3:**
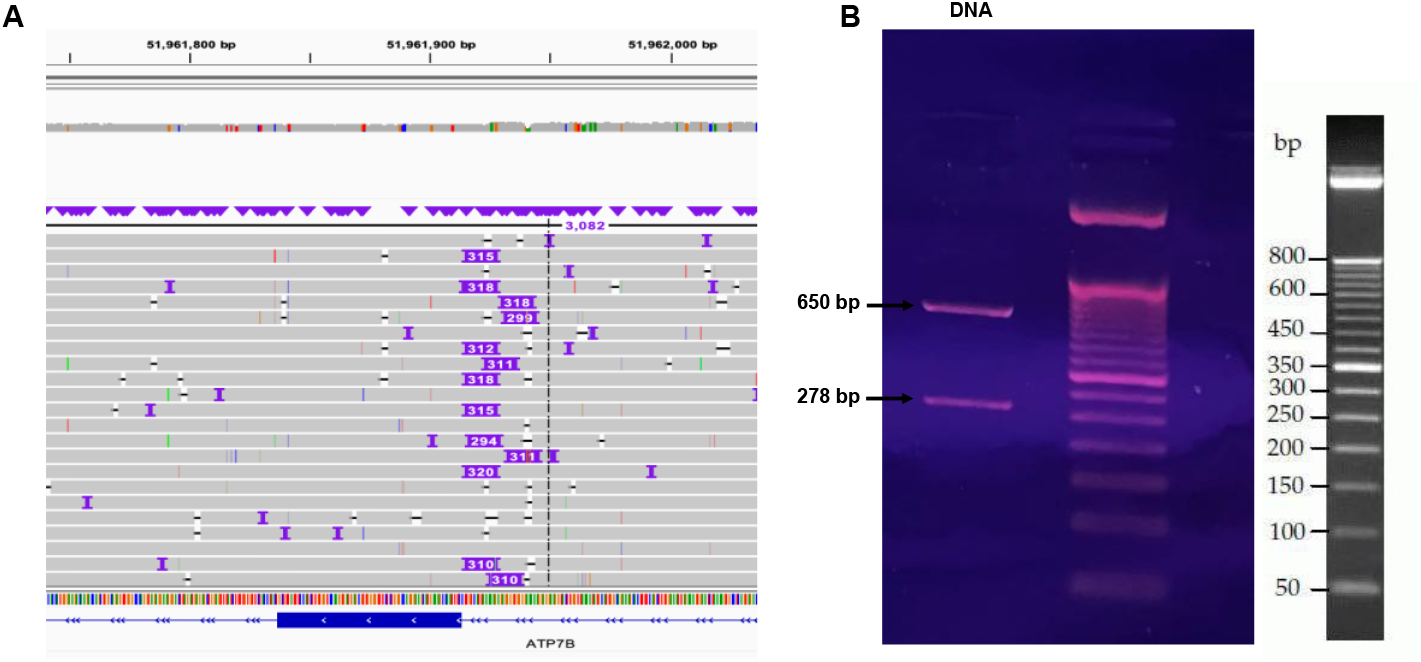
**A:** IGV visualization of the alu sequence c.1870-9_10ins at position “Chr13:51961922”. **B:** Agarose gel electrophoresis analysis showing two bands in the patient sample: one corresponding to the normal amplicon size (∼278 bp) and a second band at ∼650 bp.

The most observed genetic variant in our cohort was c.51+4A>T, which was identified in four patients. Among these, one patient carried the variant in a heterozygous state alongside the variant c.3207C>A (p.His1069Gln). This finding highlights its significance in Moroccan patients with WD. All sequencing results obtained with the ONT protocol were confirmed via traditional Sanger sequencing with segregation at the Laboratory of Medical Genetics and Oncogenetics from CHU-Fès, Morroco. This process ensured the accuracy and reliability of our findings.

In summary, in all but one patient, the likely causative variants were identified, either in the homozygous or compound heterozygous state.

## 4. Discussion

The main findings of this study were: (1) we demonstrated the feasibility and accuracy of a new ONT-based method to sequence completely the *ATPB7* gene in 15 patients diagnosed with WD; (2) we identified known and novel variants causing WD in Moroccan populations; (3) for 14 patients we identified with high likelihood the two pathogenic variants; and (4) we observed for the first time, using this approach, an intronic Alu insertion most likely causing WD in one patient.

Estimation of the prevalence of WD in Morocco was recently published (22). This study included each aspect of WD, including epidemiology, clinical manifestations, diagnosis, genetic analysis, treatment, and mortality rate. A high prevalence of WD in Morocco was detected compared with that in other countries, with a prevalence of 3.88 per 100,000, along with a pathogenic allele frequency of 0.15%. This may be attributed to the fact that 63.3% of the cases in the study were consanguineous marriages. The study also reported a mean duration of disease of 2.8 ± 1.9 years and a mortality rate of 31.9% (22). Our data set indicated that the ages of our patients confirmed with WD ranged from 5 to 30 years, with a median age at diagnosis of 15 years.

Currently, *ATP7B* is the only gene known to be associated with the development of WD. Numerous pathogenic variants have been described, each affecting the copper-transporting function of the ATP7B protein. Determining the most frequent variant by ethnicity is essential to support functional testing (23). In the Moroccan population, only two genetic studies involving small cohorts have been conducted in patients with WD. The limited accessibility of genetic testing, coupled with its high cost, has delayed the implementation of routine molecular diagnostics. Furthermore, the lack of a mutational hotspot and the high frequency of compound heterozygosity complicate genetic analysis in populations (24). For this purpose, the use of high-throughput methods such as NGS and comprehensive research with a large cohort may help to determine the disease burden in Morocco (7). To our knowledge, this study is the first conducted on a Moroccan cohort focused on sequencing the entire *ATP7B* gene, including both coding and noncoding regions.

In the present study, we identified 12 variants. Most of these variants have been reported in previous studies involving the Moroccan population, including c.3244-2A>G (25), c.3694A>C (p.Thr1232Pro) (7,22), and c.3059A>G (p.Lys1020Arg) (22,25). Additionally, we detected the variant c.3207C>A (p.His1069Gln) in one of our patients. The latter variant is among the most common in European descent populations, occurring in 15% to 63% of patients with WD (26). In one patient, no pathogenic *ATP7B* variant was identified. This observation could represent a phenocopy, reflecting locus heterogeneity or an alternative etiology that mimics the clinical and biochemical phenotype of WD (27). The c.51 + 4 A > T variant was the most frequently observed variant in our cohort and was identified in four patients who presented with hepatic manifestations. This variant is in the consensus sequence for the donor splice site of intron 1. It was previously identified in Morocco (7,22), Brazil (28), Spain (29), France (30), and Venezuela (31). A functional study demonstrated that individuals homozygous for the c.51 + 4A > T variant displayed altered splicing. In contrast, this was not observed in compound heterozygotes (32).

We detected the c.3646G>A (p.Val1216Met) variant in one patient, along with a novel insertion, c.1870-9_10ins, located within intron 5, identified as an Alu element insertion. To our knowledge, this specific variant has not been previously reported in major databases or the literature. Alu insertions are significant sources of genomic instability and can disrupt gene function through various mechanisms, such as exonization, alternative splicing, and premature transcription termination (33). It has been observed that Alu elements can integrate into transcripts, especially when positioned near splice junctions (33). We propose that the c. 1870--9_10ins Alu insertions may contribute to WD by disrupting normal RNA processing. This mechanism would be similar to the pathogenic process described in a case of WD in a 5-year-old patient from southern Italy (34). In this case, a homozygous 3039-bp deletion (c.52-2671_368del3039) from intron 1 to exon 2 of the *ATP7B* gene led to Alu exonization and abnormal splicing. The researchers identified an abnormal inclusion of cryptic splice sites derived from an Alu element, resulting in the first 577 bases of exon 2 being replaced by the exonization of a 94-base pair antisense Alu repeat. The deletion identified in their study likely produces a protein that is missing nearly the entire *ATP7B* gene, indicating that it is a severe variant associated with a severe phenotype (34). These findings underline the pathogenic potential of Alu elements in disrupting *ATP7B* function.

Despite the utility of molecular genetic testing, some patients with clinical WD do not exhibit variants within the coding regions of *ATP7B*, which could be attributed to deep intronic variants, regulatory variants, or structural variants. These findings underscore the need for more comprehensive genetic assessments (35,36). Notably, one of our patients exhibited an Alu sequence insertion in the intronic region of intron 5, further illustrating the complexity of the genetic factors involved in this condition.

To address these gaps, advanced methods such as long-read sequencing via nanopore technology combined with long-range PCR offer a comprehensive solution. This approach allows for the detection of both SNVs and SVs in a single test. Compared with traditional techniques such as Sanger sequencing and MLPA, which require multiple testing rounds, it reduces both time and cost (37,38). Additionally, ONT sequencing can identify structural rearrangements, such as inversions, that are often missed by MLPA. Investment in these emerging technologies is crucial for improving diagnostic and therapeutic options for WD in underrepresented populations.

We must recognize the limitations of this study, notably the small sample size of patients involved. Additionally, we were unable to confirm the alterations caused by the Alu insertion at the transcript level due to low expression of *ATP7b* in the blood. Furthermore, the patient declined to undergo a liver biopsy for this confirmation.

## 5. Conclusion

In conclusion, the mutational spectrum of the *ATP7B* gene in the Moroccan population is diverse and remains largely unexplored. To the best of our knowledge, this is the first study aimed at sequencing both the coding and noncoding regions of the *ATP7B* gene via ONT. The identification of the c. 1870-9_10ins Alu variant highlights the importance of sequencing the entire gene rather than focusing solely on exonic regions.

## Data availability

The datasets used and/or analysed during the current study are available from the corresponding author upon reasonable request.

## Acknowledgement

The authors thank the patients for agreeing to participate in the study, and the Interfaculty Bioinformatics Unit (IBU) for the data analysis pipeline.

## Author contributions

**Nada El Makhzen:** Conceptualisation, Methodology, Investigation, Formal analysis, Validation, Writing - Original Draft, Writing - Review & Editing, Visualisation. **Badreddine ElMakhzen:** Methodology, Validation, Formal analysis, Writing - Original Draft, Writing - Review & Editing. **Laila Bouguenouch:** Writing - Review & Editing. **Said Trhanint:** Investigation (MLPA confirmation). **Wafaa Bouzroud:** Investigation (Sanger sequencing confirmation). **Hakima Abid:** Resources (patient recruitment and clinical data collection). **Hugues Abriel:** Writing - Review & Editing.

## Ethical approval

Approved by the Ethics Biomedical Research Committee, Mohammed V University, Rabat, Morocco.

